# Association between recombinant herpes zoster vaccination and dementia risk in older adults newly admitted to post-acute and long-term care

**DOI:** 10.1101/2025.09.02.25333996

**Authors:** Kaleen N. Hayes, Daniel A. Harris, Kevin McConeghy, Lexie R. Grove, Richa Joshi, Lisa Han, H. Edward Davidson, Preeti Chachlani, Thomas A. Bayer, Mriganka Singh, Yasin Abul, Frank DeVone, Stefan Gravenstein

## Abstract

**Importance:** Ecological and observational studies have shown a protective association between herpes zoster (HZ) vaccination and dementia risk, yet many had methodological limitations or examined the live HZ vaccine that is no longer available in the US. Improved access to linked electronic health records for patients receiving post-acute care and long-term care permit robust comparisons of dementia risk in adults eligible to receive the recombinant HZ vaccine.

**Objective:** Emulate a randomized trial in observational data to estimate the association of the recombinant HZ vaccine (RZV) with incident dementia risk among older adults newly admitted for post-acute or long-term care in nursing homes (NHs).

**Design:** Retrospective cohort study with target trial emulation and the clone censor approach.

**Setting:** U.S. NHs that use PointClickCare as their electronic health record.

**Participants:** Individuals who were admitted to a NH between 01/01/2017-12/31/2022; Medicare fee-for-service beneficiaries; did not have prevalent dementia; and eligible to receive RZV as of admission.

**Exposures:** Receive one or more RZV doses within one year of admission vs. do not receive any RZV over four years of follow-up.

**Results:** We identified 509,926 eligible NH residents (mean age 79 years; 36% men). Among those alive, uncensored, and without dementia at 12 months of follow-up, 8,843 received one or more doses of RZV. Receipt of RZV within one year of NH admission was associated with a 5.8% lower absolute risk (95%CI: -3.9% to -7.5%) of newly diagnosed dementia over four years (risk ratio [RR] = 0.76 [95%CI: 0.69-0.84]; cumulative incidence in 1+ RZV vs. no RZV: 18.8% vs. 24.6%). Associations were smaller in men (RR=0.82 [95%CI: 0.68-1.01]) and those with prior live HZ vaccination (RR=0.86 [95%CI: 0.65-1.09]). Bias analyses based on two negative control outcomes (NCOs) attenuated, but did not fully explain, the main effect of RZV on dementia risk (bias-adjusted RR = 0.82 [wellness visit NCO] and RR = 0.88 [hip fracture NCO]).

**Conclusions and Relevance:** Administering RZV within 1 year of NH admission may reduce dementia risk. As RZV uptake was low overall, new NH residents would benefit from increased RZV vaccination uptake.

**KEY POINTS:** *Question:* Does the recombinant herpes zoster vaccine (RZV) reduce dementia risk among older adults?

*Findings:* In this cohort and trial emulation study of 509,926 patients newly admitted to skilled nursing or long-term care, we found that ≥1 RZV dose within a year was associated with a 24% relative and 6% absolute reduction in 4-year dementia risk. Effects were robust to bias analyses and were stronger in women and those without prior vaccination with the live HZ vaccine.

*Meaning:* Using causal inference methods, this observational study provides evidence that some cases of dementia may be prevented through vaccination with RZV.

## INTRODUCTION

Herpes zoster (HZ), also known as shingles, is a painful viral infection caused by the reactivation of the latent Varicella zoster virus (VZV). HZ is most common in adults above the age of 50 and in frail adults with immunocompromising conditions.^1,2^ Several clinical studies have linked HZ infections to increased dementia risk due to its neuroinflammatory and pro-thrombotic effects, as well as its contributions to the formation of misfolded proteins (e.g., neurofibrillary tangles and amyloid plaque accumulation).^3^ Given the efficacy and effectiveness of the recombinant zoster vaccine (RZV) at reducing HZ incidence in older adults^4,5^, there is growing interest in the vaccine’s potential to also reduce dementia incidence.

Some observational studies have found a reduced risk of dementia among those vaccinated with the live attenuated HZ vaccine, which is no longer available in the U.S. due to limited efficacy and effectiveness, and other concerns.^6–9^ Recently, authors leveraged the debut of the live HZ vaccine to conduct a “natural experiment”, finding an 20% reduced relative risk of dementia among vaccinated vs. unvaccinated adults in Wales. Other studies using similar ecological designs have corroborated these findings^7^, while a meta-analysis showed no significant benefit of the HZ vaccine on dementia risk.^10^ However, key limitations of the existing literature include the assessment of the off-market and less effective live vaccine and a reliance on ecological designs.

Our objective was to emulate a randomized trial of an RZV treatment strategy on dementia incidence among older adults who were newly admitted to post-acute and long-term care in the United States (US). Older adults admitted to nursing homes (NH) for post-acute or long-term care are at greater risk of HZ infections and cognitive decline, and admission represents a natural time for clinicians to assess vaccine eligibility and to implement an RZV vaccine treatment strategy.

## METHODS

### Study Design and Data Sources

We conducted a cohort study that aimed to emulate a randomized trial of the effects of administering at least one dose of RZV within one year of NH admission on incident dementia. The target trial framework is a pragmatic approach to estimating causal effects when a randomized trial cannot be feasibly conducted.^11^ Target trial emulation defines the parameters of a hypothetical and “ideal” randomized trial, and emulates those parameters as closely as possible using observational data (**eTable 1**).^11,12^ We published an a priori protocol prior to beginning analyses,^13^ and we followed the Strengthening the Reporting of Observational Studies in Epidemiology (STROBE) Guidelines for cohort studies.^14^

We used observational data from a large NH electronic health record (EHR) vendor (PointClickCare [PCC]) of community nursing homes that provide post-acute and long-term care. These data come from over 11,000 unique nursing homes with geographical representation in nearly all 50 US states. The NH EHR data comprised resident demographic information; Minimum Dataset (MDS) records, which are mandatory clinical and functional assessments completed upon nursing home admission and at least quarterly thereafter; prescription and non-prescription medication orders and administrations; vaccination history, orders, and administrations; and other elements, including vital signs. The EHR data contained information on all residents admitted to a NH that used PointClickCare between January 1, 2017, to December 31, 2022.

To measure outcome events and other clinical information that occurred prior to and following NH admission, we linked residents’ EHR data to the following Medicare claims files: Medicare Beneficiary Summary File (MBSF); Chronic Conditions Warehouse (CCW); Medicare Provider Analysis and Review (MedPAR); Medicare Part B carrier claims; and Medicare Part D medication and vaccine claims. Details and results of the linkage, as well as descriptions of the Medicare databases, are available in **eMethods 1**. The Brown University Institutional Review Board approved this study, and informed consent was waived due to the use of deidentified data.

### Population

We identified all new admissions to a NH among those successfully linked to a Medicare beneficiary between January 1, 2017 and December 31, 2022. These residents represented a mix of post-acute skilled nursing and long-term residents. We applied all eligibility criteria as of the study index date (time zero), which was defined as the first MDS admission assessment date. We excluded residents <66 years of age or who had fewer than 12 months of continuous enrollment in Medicare Parts A, B, and D prior to the index date. We also excluded residents with existing dementia; who were actively under or recently transferred from hospice; had a diagnosis of HZ in the prior six months; had previously received one or more doses of RZV; or were dead as of the index date but still present in the EHR and/or claims data. We did not exclude residents who previously received the live attenuated HZ vaccine, as U.S. guidelines^15^ recommend re-vaccination with RZV. Definitions for exclusion criteria are provided in **eTable 2**.

### Exposures and Outcomes

The treatment strategies (i.e., exposures) of interest were: 1) receive at least one dose of RZV within the first 12 months of admission and 2) never receive RZV during follow-up. We identified RZV administrations using Medicare Part D claims, a vaccine administration record in the EHR, or a new vaccine immunization record for RZV in the EHR.

We defined the outcome of incident dementia as: 1) an inpatient hospitalization with a dementia-related diagnosis code; or 2) three Part B carrier claims with a dementia-related diagnosis, separated by at least 30 days; or 3) claim or recorded administration for a dementia medication (i.e., acetylcholinesterase inhibitor or memantine; see **eTable 2** for detailed variable definitions). This outcome definition was estimated to have a sensitivity of 79.3% and specificity of 99.1% in community-dwelling older adults.^16^

### Covariates

We measured demographic and clinical characteristics associated with dementia and receiving RZV based on prior literature and clinical rationale. Demographics were measured via the MBSF (e.g., age, race and ethnicity), and clinical characteristics (e.g., Cognitive Function Scale^17^, healthcare utilization, prior stroke) were measured using MDS assessments combined with CCW indicators as well as Parts A (from MedPAR), B, and D claims (**eTable 2**). Select covariates were updated over time using 30-day lookback windows (e.g., time-varying indicators for recently administered vaccinations, HZ infections, and health services use).

### Clone-Censor-Weighting and Follow-up

We utilized the “clone-censor-weight” approach in the target trial emulation.^18,19^ Details of this method are provided in **eMethods 2**. In brief, all eligible new residents were “cloned” (copied), and assigned to one of the two treatment strategies. The first 12 months of follow-up were considered a “grace period” to allow for clones who were assigned to vaccination to receive at least one dose of RZV. Clones were followed until they experienced the outcome, were artificially censored due to non-adherence to their assigned treatment strategy, or experienced a true censoring event (death, entry to hospice, disenrollment from Medicare, 4 years after the index date; or end of data [December 31, 2022]). A study figure is presented in **eFigure 1**.

**Figure 1.**
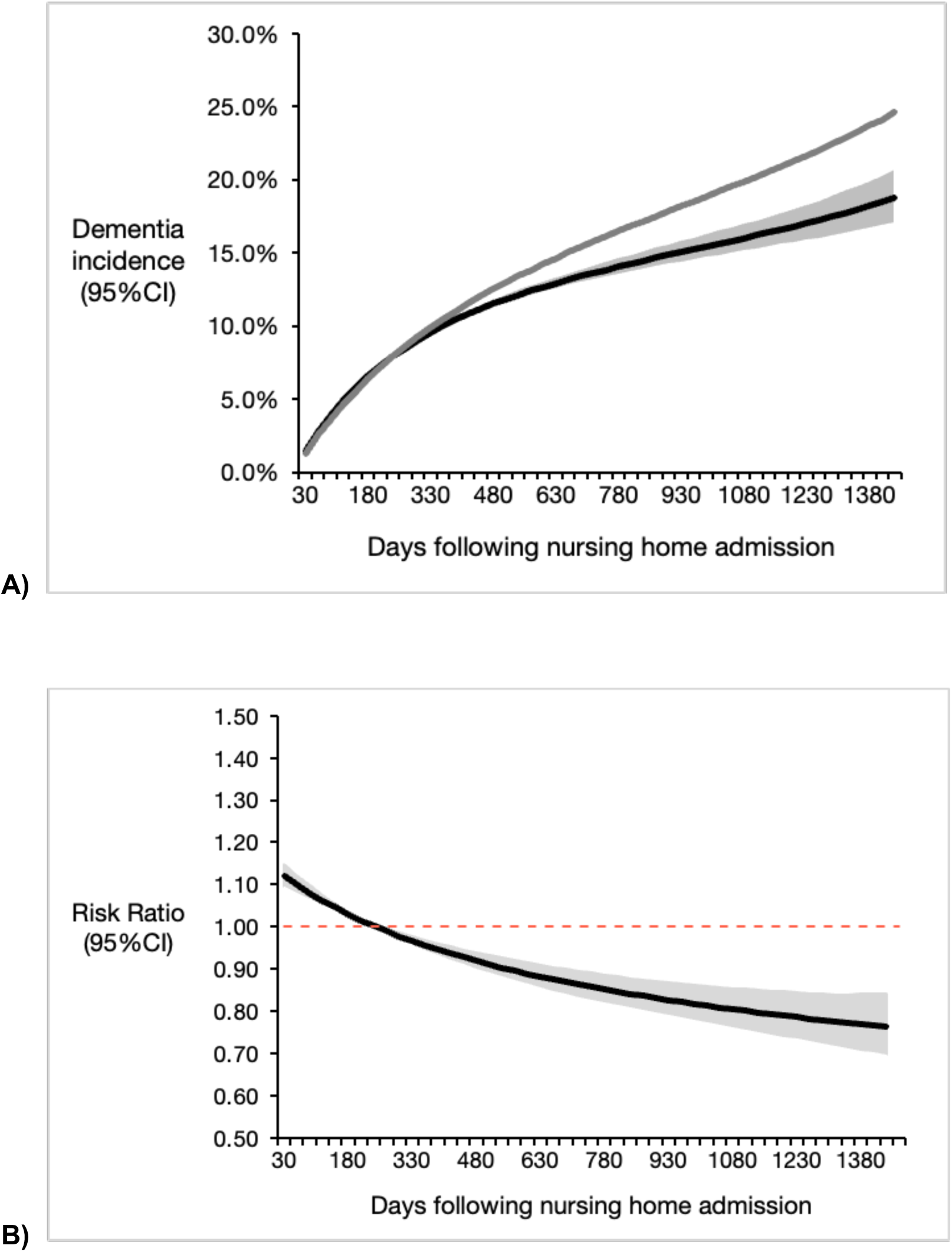
A) Adjusted cumulative incidence of dementia by RZV treatment strategy and B) relative risk of dementia over 4 years of follow-up. *NB: In Panel A, the black line indicates receipt of 1+ RZV dose, and the gray line indicates no RZV receipt. Shaded regions indicate 95% confidence intervals generated using percentile-based Poisson bootstrapping with 500 replicates*.

### Statistical Analyses

We estimated the inverse probability of remaining uncensored (IPCW) for each treatment strategy using pooled logistic regression with baseline and time-varying covariates (**eMethods 2**).^20,21^ Pooled logistic regression with IPCW compared the cumulative incidence of dementia in 30-day intervals between the RZV treatment groups. To assess the impact of different covariates on the point estimates, we calculated unadjusted, baseline-adjusted, and baseline and time-varying adjusted IPCW models. Percentile-based Poisson bootstrapping with 500 replicates was used to generate 95% confidence intervals for all estimates.

### Subgroups and Sensitivity Analyses

We repeated the primary analysis in *a priori* defined subgroups stratified by sex and prior receipt of the live HZ vaccine. We then conducted several sensitivity analyses to assess the robustness of our findings:

First, we explored the impact of potential outcome misclassification by implementing more sensitive and specific definitions of dementia. The sensitive definition was defined as any inpatient or outpatient claim with a dementia code, or new use of a dementia medication. The specific definition was defined as new use of a dementia medication only. Second, we conducted a negative control outcome (NCO) analysis using hip fracture and wellness outpatient visits to explore the extent of residual bias.^22^ In theory, RZV is not causally associated with the NCOs, but they share similar confounding structures as the primary RZV and dementia association of interest. The presence of an adjusted association between the treatment strategies and NCOs suggests residual bias and can facilitate calibration of the primary findings through quantitative bias analysis. While some studies only consider there to be meaningful residual confounding if the NCO remains statistically significant after full adjustment, we calibrated our primary risk ratio of the RZV-dementia association according to the strength of an unmeasured continuous confounder that would nullify the NCO’s point estimate. In other words, we determined the strengths of association between an unmeasured continuous confounder, RZV, and NCO to fully attenuate the RZV-NCO risk ratio to 1.0 (i.e., the RZV-NCO association is hypothetically null [RR=1.0] under all necessary assumptions).^23^ Third, we modified follow-up assumptions and model specifications by (1) exploring the potential competing risk of death via a multinomial logistic regression model with dementia and death as joint outcomes; (2) censoring residents on NH discharge; (3) truncating IPCW at the 99th percentile; (4) stabilizing IPCW according to the cumulative probability of vaccination in each interval; and (5) adding all baseline covariates to the dementia outcome model.

## RESULTS

### Study Populations

After linkage and applying eligibility criteria, we identified 509,926 residents (average age=79.3 [SD=8.2] years; 63.6% women [**Table 1**; see **eTable 3** for exclusions flow]). Most residents were short stay (median number of days until discharge = 18 days [Q1=9, Q3=36]), though the length of stay was right-skewed (mean=44 days [SD=127]).

**Table 1.**
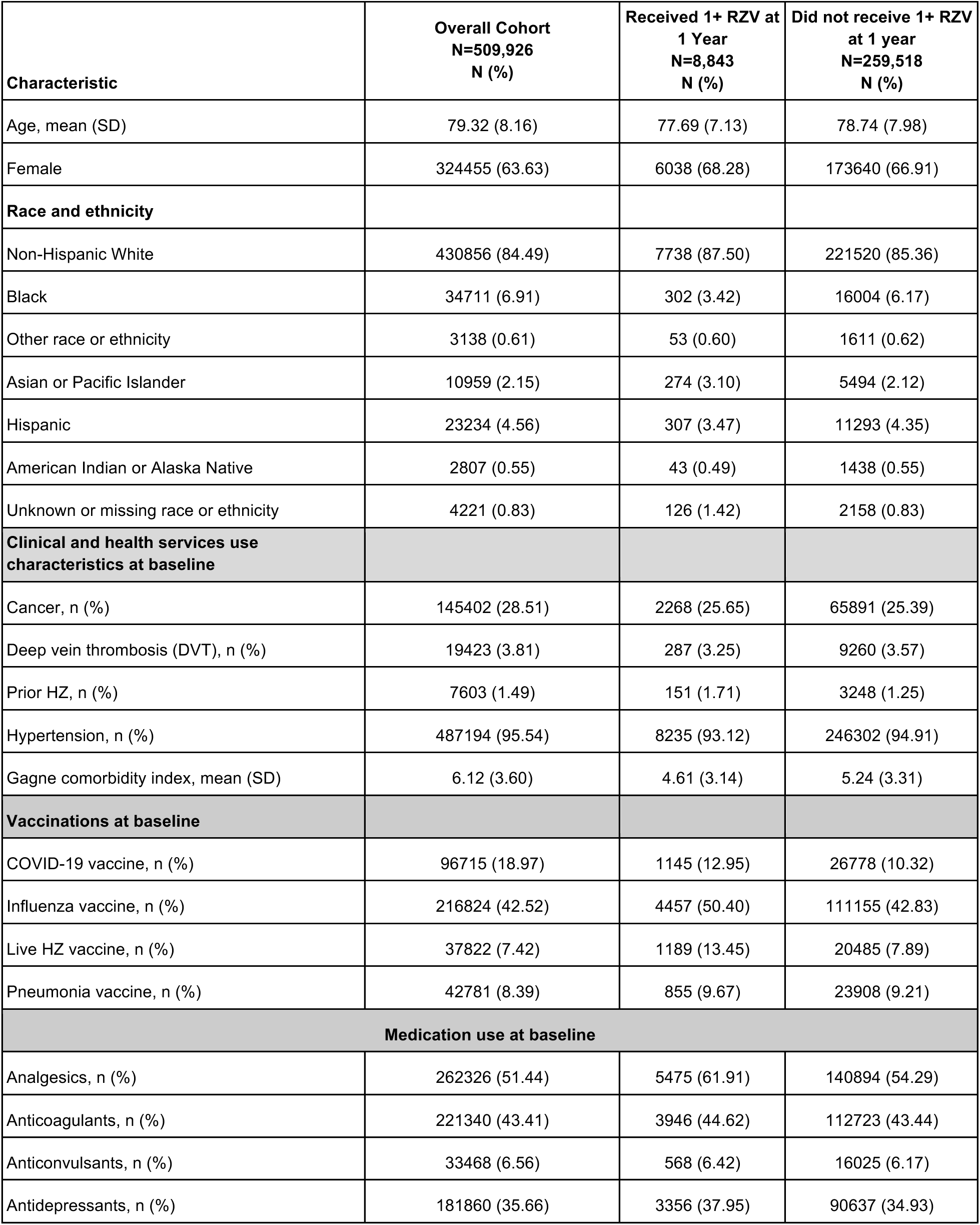
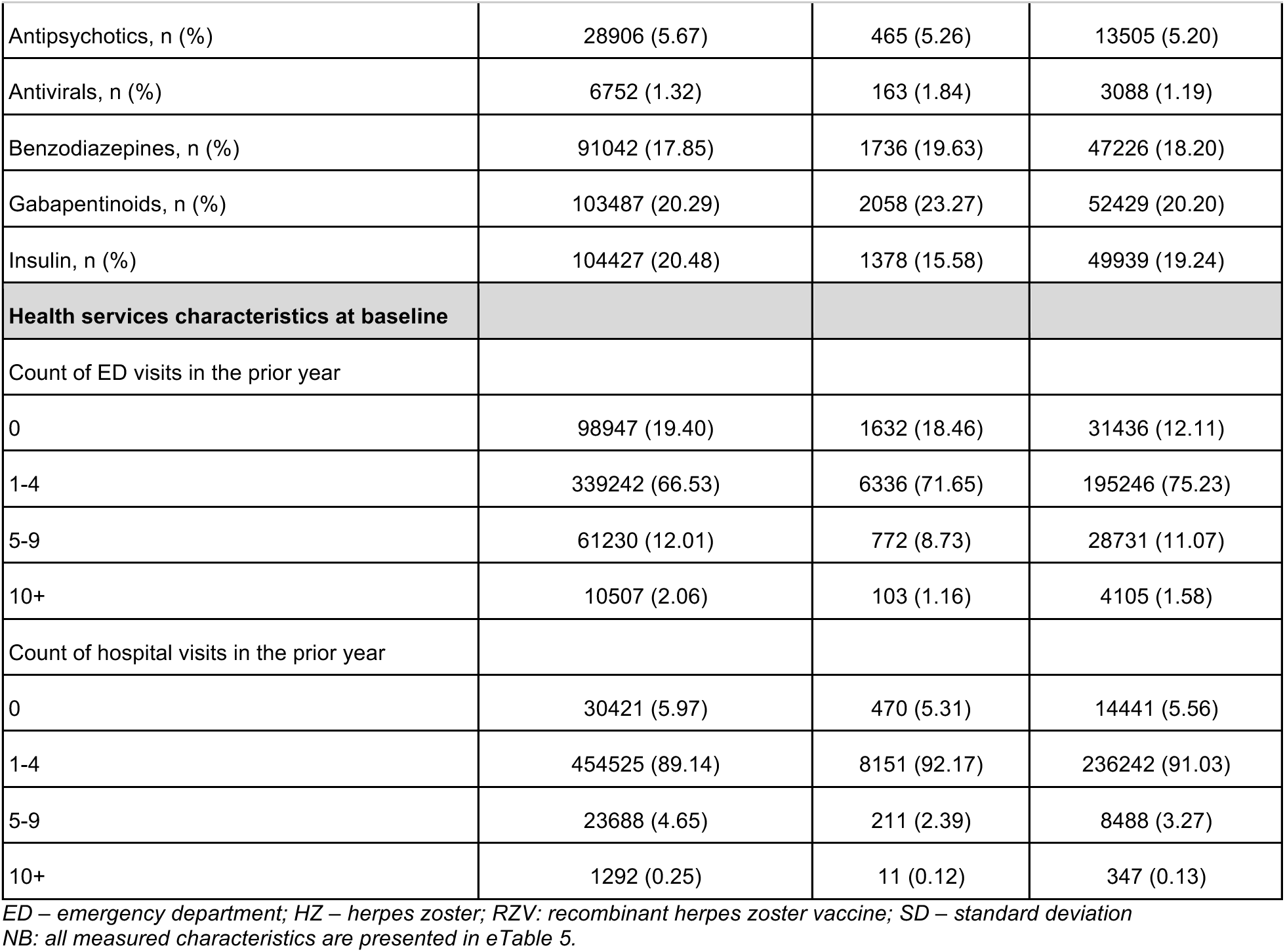
Select demographic and clinical characteristics of nursing home residents eligible for the target trial at admission and at one year following admission.

| Characteristic | Overall Cohort<br>N=509,926<br>N (%) | Received 1+ RZV at<br>1 Year<br>N=8,843<br>N (%) | Did not receive 1+ RZV<br>at 1 year<br>N=259,518<br>N (%) |
| --- | --- | --- | --- |
| Age, mean (SD) | 79.32 (8.16) | 77.69 (7.13) | 78.74 (7.98) |
| Female | 324455 (63.63) | 6038 (68.28) | 173640 (66.91) |
| **Race and ethnicity** |  |  |  |
| Non-Hispanic White | 430856 (84.49) | 7738 (87.50) | 221520 (85.36) |
| Black | 34711 (6.91) | 302 (3.42) | 16004 (6.17) |
| Other race or ethnicity | 3138 (0.61) | 53 (0.60) | 1611 (0.62) |
| Asian or Pacific Islander | 10959 (2.15) | 274 (3.10) | 5494 (2.12) |
| Hispanic | 23234 (4.56) | 307 (3.47) | 11293 (4.35) |
| American Indian or Alaska Native | 2807 (0.55) | 43 (0.49) | 1438 (0.55) |
| Unknown or missing race or ethnicity | 4221 (0.83) | 126 (1.42) | 2158 (0.83) |
| **Clinical and health services use characteristics at baseline** |  |  |  |
| Cancer, n (%) | 145402 (28.51) | 2268 (25.65) | 65891 (25.39) |
| Deep vein thrombosis (DVT), n (%) | 19423 (3.81) | 287 (3.25) | 9260 (3.57) |
| Prior HZ, n (%) | 7603 (1.49) | 151 (1.71) | 3248 (1.25) |
| Hypertension, n (%) | 487194 (95.54) | 8235 (93.12) | 246302 (94.91) |
| Gagne comorbidity index, mean (SD) | 6.12 (3.60) | 4.61 (3.14) | 5.24 (3.31) |
| **Vaccinations at baseline** |  |  |  |
| COVID-19 vaccine, n (%) | 96715 (18.97) | 1145 (12.95) | 26778 (10.32) |
| Influenza vaccine, n (%) | 216824 (42.52) | 4457 (50.40) | 111155 (42.83) |
| Live HZ vaccine, n (%) | 37822 (7.42) | 1189 (13.45) | 20485 (7.89) |
| Pneumonia vaccine, n (%) | 42781 (8.39) | 855 (9.67) | 23908 (9.21) |
| **Medication use at baseline** |  |  |  |
| Analgesics, n (%) | 262326 (51.44) | 5475 (61.91) | 140894 (54.29) |
| Anticoagulants, n (%) | 221340 (43.41) | 3946 (44.62) | 112723 (43.44) |
| Anticonvulsants, n (%) | 33468 (6.56) | 568 (6.42) | 16025 (6.17) |
| Antidepressants, n (%) | 181860 (35.66) | 3356 (37.95) | 90637 (34.93) |
| Antipsychotics, n (%) | 28906 (5.67) | 465 (5.26) | 13505 (5.20) |
| Antivirals, n (%) | 6752 (1.32) | 163 (1.84) | 3088 (1.19) |
| Benzodiazepines, n (%) | 91042 (17.85) | 1736 (19.63) | 47226 (18.20) |
| Gabapentinoids, n (%) | 103487 (20.29) | 2058 (23.27) | 52429 (20.20) |
| Insulin, n (%) | 104427 (20.48) | 1378 (15.58) | 49939 (19.24) |
| **Health services characteristics at baseline** |  |  |  |
| Count of ED visits in the prior year |  |  |  |
| 0 | 98947 (19.40) | 1632 (18.46) | 31436 (12.11) |
| 1-4 | 339242 (66.53) | 6336 (71.65) | 195246 (75.23) |
| 5-9 | 61230 (12.01) | 772 (8.73) | 28731 (11.07) |
| 10+ | 10507 (2.06) | 103 (1.16) | 4105 (1.58) |
| Count of hospital visits in the prior year |  |  |  |
| 0 | 30421 (5.97) | 470 (5.31) | 14441 (5.56) |
| 1-4 | 454525 (89.14) | 8151 (92.17) | 236242 (91.03) |
| 5-9 | 23688 (4.65) | 211 (2.39) | 8488 (3.27) |
| 10+ | 1292 (0.25) | 11 (0.12) | 347 (0.13) |

ED – emergency department; HZ – herpes zoster; RZV: recombinant herpes zoster vaccine; SD – standard deviation
NB: all measured characteristics are presented in eTable 5.

In the first 12 months of follow-up, 7.2% were diagnosed with dementia; 12% died; and 28% disenrolled, entered hospice, or reached the end of available data (reasons for censoring in **eTable 4**). Among those alive, uncensored, and without dementia by month 12 of follow-up, 8,843 clones adhered to the treatment strategy to receive one or more doses of RZV within a year of nursing home admission (of which 68.73% had evidence of a second RZV dose), and 259,518 remained unvaccinated. Compared to those who did not receive any RZV, vaccinated individuals were younger (average age 77.7 vs. 78.7 years), less likely to be of Black or Hispanic race (e.g., 3% vs. 6% Black race), and had fewer comorbidities at baseline (e.g., average combined comorbidity score 4.6 vs. 5.2) (**Table 1** and **eTable 5**). Those who received RZV also were more likely to have received the live HZ vaccine prior (13% vs. 8%).

### Censoring Weighting and Distribution

Among the 57 baseline and time-varying covariates included in the IPCW models, prior receipt of the live HZ vaccine; influenza vaccine; prior HZ diagnosis; year of admission to the NH; and discharging from the NH were associated with receiving RZV within 12 months (**eTable 6**). Older age, a higher comorbidity burden, and greater healthcare utilization were associated with a lower likelihood of RZV. IPCW were reasonably distributed (**eTable 7**), with a median weight of 1.0 in both treatment groups.

### Incidence of Dementia Between Vaccination Groups

The unadjusted cumulative incidence of dementia over the 4-year follow-up was 18.3% in the RZV treatment group and 24.8% in the no RZV group (unadjusted RR=0.74 [95%CI 0.66 to 0.81]; RD=-6.5% [95%CI -8.1% to -4.7%]). In pooled models applying IPCW containing both baseline and time-varying covariates, receiving RZV in the first year of nursing home admission was associated with a lower risk of dementia (Relative risk [RR]=0.76 [95%CI 0.69 to 0.84]; Risk difference [RD]= -5.8% [95%CI -7.5% to -3.9%]) (**Table 2**; **Figure 1**). Estimates and number of events by year of follow-up are presented in **eTable 8**.

**Table 2.**
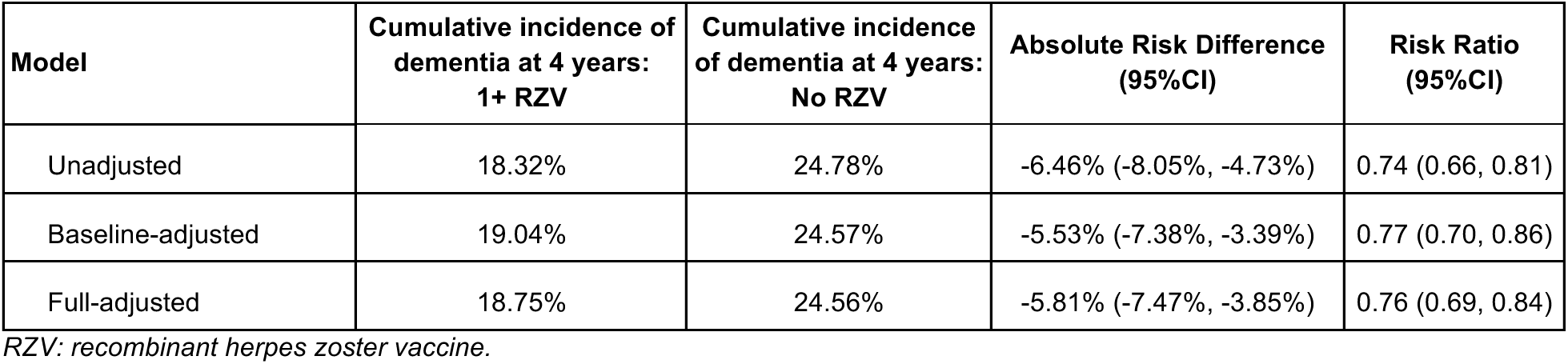
Absolute and relative risk of dementia between residents who received at least one dose of RZV within one year of admission compared to no RZV at four years of follow-up.

The association between RZV and dementia was attenuated in men vs. women (RR=0.82 [95% CI 0.68 to 1.01] vs. RR=0.75 [95%CI 0.67 to 0.84]), and among those with prior receipt of the live HZ vaccine (RR=0.86 [95%CI 0.65 to 1.09]) vs. those without (RR=0.75 [95%CI 0.67 to 0.84]) (**Figure 2**).

**Figure 2.**
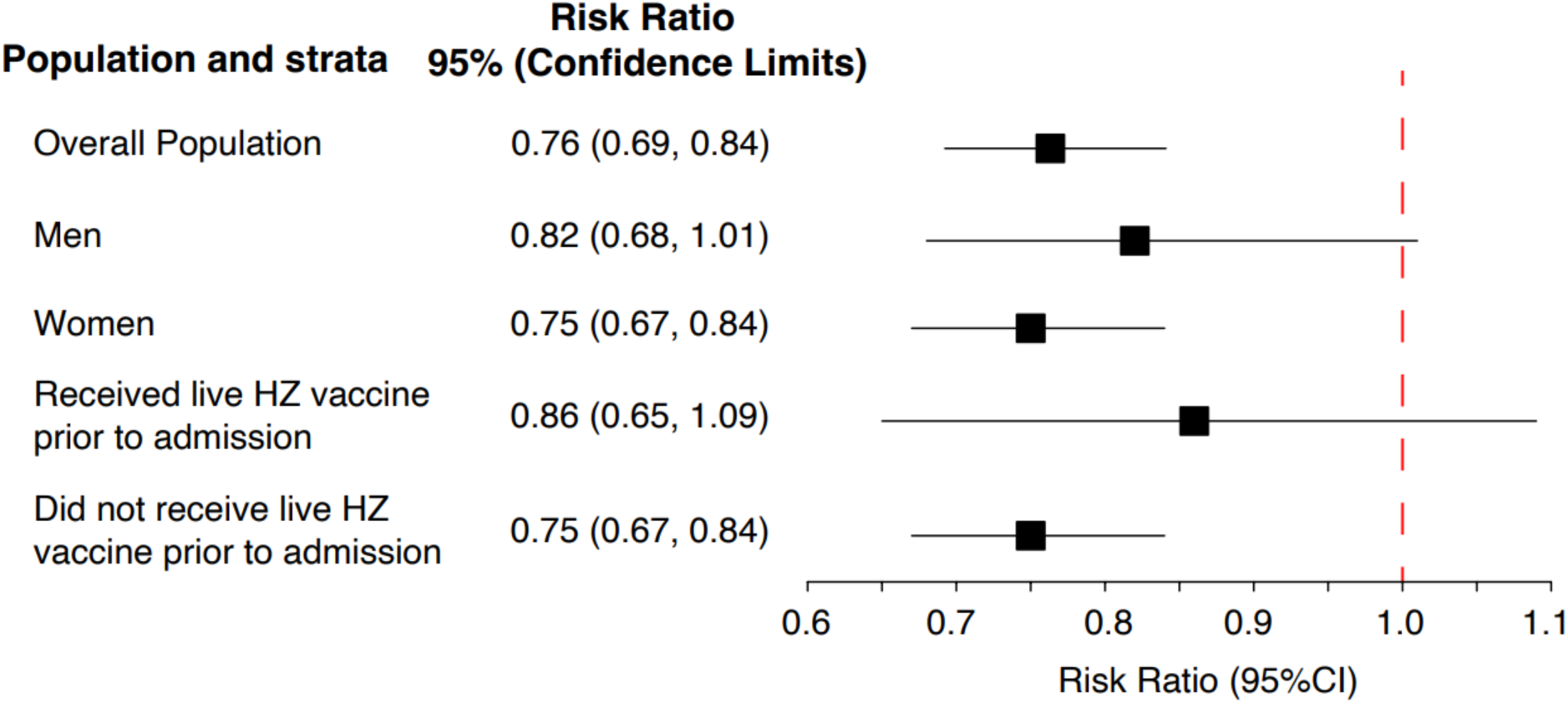
Relative risk of dementia between residents who received at least one dose of RZV within one year of admission compared to no RZV at four years of follow-up, stratified by sex and prior receipt of the live HZ vaccine.

### Sensitivity Analyses

Unadjusted and fully adjusted results when modifying the dementia outcome definition are available in **eTable 9**. When broadening the definition of the dementia outcome to any diagnostic code or new anti-dementia medication use, results were consistent with the primary analysis; however, the specific outcome of receiving a new dementia medication alone was attenuated (RR=0.93 [95%CI 0.76 to 1.11]). Next, associations between RZV and both NCOs became statistically insignificant after full adjustment (**eTable 10**). Using bias parameters to nullify the NCOs point estimates to 1.0 attenuated the association between RZV and dementia to RR=0.88, for hip fracture bias correction, and to RR=0.82 for wellness visit bias correction. Fully adjusted multinomial models accounting for the competing risk of death did not meaningfully change the main findings, though we found that those who received RZV had a lower risk of death (RR=0.76 [95% CI 0.75 to 0.76]). Censoring on NH discharge, and truncating and stabilizing the IPCW did not meaningfully alter results (**eTable 11**).

## DISCUSSION

We emulated a randomized trial in a cohort of over 500,000 older adults with a recent admission to a NH and used causal inference methods to estimate the association between receiving at least 1 RZV dose within a year and incident dementia. We found that those who received RZV had a 24% reduced relative risk of new dementia and a 6% reduction in absolute risk after 4 years compared to those who did not receive the vaccine. These estimates translate to one fewer dementia case per 17 older adults entering a NH setting who receive RZV. Negative control outcomes became statistically insignificant after full IPCW adjustment, though suggest residual bias in the fully-adjusted point estimates; however, calibrating our primary RZV-dementia estimates according to this potential bias did not explain the observed protective benefit of RZV on dementia risk.

To our knowledge, this is the largest person-level observational study of older adults and RZV to date and addresses a gap in knowledge regarding the potential effects of vaccination against HZ on dementia risk. Several recent studies have leveraged quasi-experimental methods to examine associations between the live HZ vaccine and dementia and found protective effects. In the last year, Eyting and Pomirchy separately examined the probability of dementia diagnosis among those eligible vs. not eligible for the live HZ vaccine in Wales and Australia, respectively, using regression discontinuity designs.^7,8^ Both studies found a significant reduction in the risk of dementia (3.5% absolute reduction in Wales and 1.5% absolute reduction in Australia, each over around 7 years’ follow-up). Prior to these studies, other epidemiologic examinations on RZV^24–30^ found a protective association of HZ vaccination with dementia of similar magnitude to our estimates, though many mixed RZV and the live attenuated vaccine exposures. Our causal inference study emulating a target trial in U.S. older adults admitted to a NH corroborates and extends prior work by demonstrating a significant association with dementia risk reduction with RZV.

The mechanism underlying the potential protective effect of HZ vaccination is likely multifold and complex. Dementia is a heterogenous condition, of which half have a mixed-type etiology.^31^ VZV is a neuroinflammatory virus that directly damages cerebral arteries, reduces blood flow through inflammation, and increases the risk of cerebrovascular events, all phenomena that potentially contribute to vascular dementia.^3,32^ It has been posited that HZ prevention through vaccination can eliminate or delay the virus’s inflammatory cascade, reducing cytokine release, microglial activation, and stroke — events that independently and jointly contribute to cognitive impairment and dementia.^33^ Further, *in vitro* evidence suggests herpesviruses may contribute to Alzheimer’s Disease and Lewy Body dementias through increasing formation and accumulation of misfolded oligomers, amyloid plaques, and neurofibrillary tangles.^34–36^ Finally, vaccination in general may provide protective immunomodulatory effects discussed in detail elsewhere.^37,38^

Subgroup analyses from our study corroborate some of these potential biological mechanisms and generate additional hypotheses. First, we observed that the receipt of RZV in addition to the live HZ vaccine had attenuated benefits (i.e., 14% reduction in relative risk which was not statistically significant vs. 25% for those without prior live vaccine receipt). RZV also appeared to have an attenuated effect in men vs. women. Research consistently shows that women experience higher immunogenicity and effectiveness, as well as more adverse vaccine events, than men, which may explain this difference between sexes.^38^

Key strengths of this study include its large sample of U.S. NH residents, comprehensive capture of vaccine exposures, dementia outcomes, and time-varying covariates with linked EHR data, and causal inference framework to address immortal time bias. However, there are several limitations. First, point estimates from the negative control outcomes suggest that residual bias explains some, but not all, of the protective effect of RZV on dementia risk. Second, follow-up time was limited due to the approval of RZV in 2017 and the extent of available EHR and claims data. Third, measurement of RZV and dementia were imperfect due to incomplete capture of vaccines administered prior to Part D enrollment and the reliance on administrative data algorithms for dementia. We also were unable to explore effects on type of dementia given the nonspecific nature of administrative data for capturing dementia subtypes and rarity of diagnosing the full etiology in routine clinical practice.

## Conclusions

Using observational data to emulate a randomized trial, we found a 24% relative risk reduction in new dementia diagnosis among U.S. older adults recently admitted to a NH who received at least one dose of RZV. Though the observational evidence is robust and largely consistent, randomized trials are needed to confirm the causal association of the recombinant HZ vaccination and dementia risk. A pragmatic, randomized trial of NH residents may be a feasible step to establish more definitive causal evidence in this population due to the wealth of clinical and functional data that are regularly collected in this setting. Nevertheless, given the evidence for effectiveness of RZV to reduce HZ infections, RZV is a feasible and cost-effective preventative intervention for older adults to reduce comorbidity and improve quality of life.

## Data Availability

Data Sharing Statement: These data were obtained through data use agreements with the Centers for Medicare and Medicaid Services and PointClickCare(R). Statistical code related to clone censor weighting is available at https://kmcconeghy.github.io/ccwtutorial/. Other statistical code may be provided upon reasonable request.

## Data Sharing Statement

These data were obtained through data use agreements with the Centers for Medicare and Medicaid Services and PointClickCare(R) that prohibit distribution. Statistical code related to clone censor weighting is available at https://kmcconeghy.github.io/ccwtutorial/. Other statistical code may be provided upon reasonable request.

## REFERENCES

1. John AR, Canaday DH. Herpes Zoster in the Older Adult. Infect Dis Clin North Am. 2017;31(4):811–826. doi:10.1016/j.idc.2017.07.016

2. Marra F, Parhar K, Huang B, Vadlamudi N. Risk Factors for Herpes Zoster Infection: A Meta-Analysis. Open Forum Infect Dis. 2020;7(1):ofaa005. doi:10.1093/ofid/ofaa005

3. Elhalag RH, Motawea KR, Talat NE, et al. Herpes Zoster virus infection and the risk of developing dementia: A systematic review and meta-analysis. Medicine (Baltimore). 2023;102(43):e34503. doi:10.1097/MD.0000000000034503

4. Cunningham AL, Lal H, Kovac M, et al. Efficacy of the Herpes Zoster Subunit Vaccine in Adults 70 Years of Age or Older. N Engl J Med. 2016;375(11):1019–1032. doi:10.1056/NEJMoa1603800

5. Tseng HF, Sy LS, Ackerson BK, et al. Effectiveness of the Adjuvanted Recombinant Zoster Vaccine in Adults ≥50 Years in the United States. Clin Infect Dis. Published online June 23, 2025:ciaf329. doi:10.1093/cid/ciaf329

6. Lehrer S, Rheinstein PH. Herpes Zoster Vaccination Reduces Risk of Dementia. In Vivo. 2021;35(6):3271–3275. doi:10.21873/invivo.12622

7. Pomirchy M, Bommer C, Pradella F, Michalik F, Peters R, Geldsetzer P. Herpes Zoster Vaccination and Dementia Occurrence. JAMA. Published online April 23, 2025. doi:10.1001/jama.2025.5013

8. Eyting M, Xie M, Michalik F, Heß S, Chung S, Geldsetzer P. A natural experiment on the effect of herpes zoster vaccination on dementia. Nature. Published online April 2, 2025. doi:10.1038/s41586-025-08800-x

9. Update on Herpes Zoster Vaccine: Licensure for Persons Aged 50 Through 59 Years. Centers for Disease Control and Prevention. November 11, 2011. Accessed August 19, 2025. https://www.cdc.gov/mmwr/preview/mmwrhtml/mm6044a5.htm

10. Shah S, Dahal K, Thapa S, et al. Herpes zoster vaccination and the risk of dementia: A systematic review and meta-analysis. Brain Behav. 2024;14(2):e3415. doi:10.1002/brb3.3415

11. Hernán MA, Wang W, Leaf DE. Target Trial Emulation: A Framework for Causal Inference From Observational Data. JAMA. 2022;328(24):2446–2447. doi:10.1001/jama.2022.21383

12. Hernán MA, Robins JM. Using Big Data to Emulate a Target Trial When a Randomized Trial Is Not Available. Am J Epidemiol. 2016;183(8):758–764. doi:10.1093/aje/kwv254

13. Harris D, Abul Y, Hayes K, et al. Estimating the effect of herpes zoster vaccination on dementia risk: a target trial approach in US nursing homes. Published online April 30, 2024. Accessed August 6, 2025. https://osf.io/e67r8/

14. von Elm E, Altman DG, Egger M, et al. The Strengthening the Reporting of Observational Studies in Epidemiology (STROBE) statement: guidelines for reporting observational studies. Ann Intern Med. 2007;147(8):573–577. doi:10.7326/0003-4819-147-8-200710160-00010

15. Anderson TC, Masters NB, Guo A, et al. Use of Recombinant Zoster Vaccine in Immunocompromised Adults Aged ≥19 Years: Recommendations of the Advisory Committee on Immunization Practices — United States, 2022. MMWR Morb Mortal Wkly Rep. 2022;71(3):80–84. doi:10.15585/mmwr.mm7103a2

16. Jaakkimainen RL, Bronskill SE, Tierney MC, et al. Identification of Physician-Diagnosed Alzheimer’s Disease and Related Dementias in Population-Based Administrative Data: A Validation Study Using Family Physicians’ Electronic Medical Records. JAD. 2016;54(1):337–349. doi:10.3233/JAD-160105

17. Thomas KS, Dosa D, Wysocki A, Mor V. The Minimum Data Set 3.0 Cognitive Function Scale. Med Care. 2017;55(9):e68–e72. doi:10.1097/MLR.0000000000000334

18. Gaber CE, Ghazarian AA, Strassle PD, et al. De-Mystifying the Clone-Censor-Weight Method for Causal Research Using Observational Data: A Primer for Cancer Researchers. Cancer Med. 2024;13(23):e70461. doi:10.1002/cam4.70461

19. Hernán MA, Sauer BC, Hernández-Díaz S, Platt R, Shrier I. Specifying a target trial prevents immortal time bias and other self-inflicted injuries in observational analyses. J Clin Epidemiol. 2016;79:70–75. doi:10.1016/j.jclinepi.2016.04.014

20. Zhao SS, Lyu H, Yoshida K. Versatility of the clone-censor-weight approach: response to “trial emulation in the presence of immortal-time bias.” International Journal of Epidemiology. 2021;50(2):694–695. doi:10.1093/ije/dyaa223

21. Duchesneau ED, Jackson BE, Webster-Clark M, et al. The Timing, the Treatment, the Question: Comparison of Epidemiologic Approaches to Minimize Immortal Time Bias in Real-World Data Using a Surgical Oncology Example. Cancer Epidemiol Biomarkers Prev. 2022;31(11):2079–2086. doi:10.1158/1055-9965.EPI-22-0495

22. Lipsitch M, Tchetgen Tchetgen E, Cohen T. Negative controls: a tool for detecting confounding and bias in observational studies. Epidemiology. 2010;21(3):383–388. doi:10.1097/EDE.0b013e3181d61eeb

23. McGowan LD. tipr: An R package for sensitivity analyses forunmeasured confounders. JOSS. 2022;7(77):4495. doi:10.21105/joss.04495

24. Scherrer JF, Salas J, Wiemken TL, Hoft DF, Jacobs C, Morley JE. Impact of herpes zoster vaccination on incident dementia: A retrospective study in two patient cohorts. PLoS One. 2021;16(11):e0257405. doi:10.1371/journal.pone.0257405

25. Harris K, Ling Y, Bukhbinder AS, et al. The Impact of Routine Vaccinations on Alzheimer’s Disease Risk in Persons 65 Years and Older: A Claims-Based Cohort Study using Propensity Score Matching. JAD. 2023;95(2):703–718. doi:10.3233/JAD-221231

26. Lophatananon A, Mekli K, Cant R, et al. Shingles, Zostavax vaccination and risk of developing dementia: a nested case–control study—results from the UK Biobank cohort. BMJ Open. 2021;11(10):e045871. doi:10.1136/bmjopen-2020-045871

27. Tang E, Ray I, Arnold BF, Acharya NR. Recombinant zoster vaccine and the risk of dementia. Vaccine. 2025;46:126673. doi:10.1016/j.vaccine.2024.126673

28. Schnier C, Janbek J, Lathe R, Haas J. Reduced dementia incidence after varicella zoster vaccination in Wales 2013-2020. Alzheimers Dement (N Y). 2022;8(1):e12293. doi:10.1002/trc2.12293

29. Wilkinson T, Schnier C, Bush K, et al. Drug prescriptions and dementia incidence: a medication-wide association study of 17000 dementia cases among half a million participants. J Epidemiol Community Health. 2022;76(3):223–229. doi:10.1136/jech-2021-217090

30. Ukraintseva S, Yashkin AP, Akushevich I, et al. Associations of infections and vaccines with Alzheimer’s disease point to a role of compromised immunity rather than specific pathogen in AD. Experimental Gerontology. 2024;190:112411. doi:10.1016/j.exger.2024.112411

31. James BD, Wilson RS, Boyle PA, Trojanowski JQ, Bennett DA, Schneider JA. TDP-43 stage, mixed pathologies, and clinical Alzheimer’s-type dementia. Brain. 2016;139(11):2983–2993. doi:10.1093/brain/aww224

32. Nagel MA, Cohrs RJ, Mahalingam R, et al. The varicella zoster virus vasculopathies: Clinical, CSF, imaging, and virologic features. Neurology. 2008;70(11):853–860. doi:10.1212/01.wnl.0000304747.38502.e8

33. Cairns DM, Itzhaki RF, Kaplan DL. Potential Involvement of Varicella Zoster Virus in Alzheimer’s Disease via Reactivation of Quiescent Herpes Simplex Virus Type 1. JAD. 2022;88(3):1189–1200. doi:10.3233/JAD-220287

34. Álvarez G, Aldudo J, Alonso M, Santana S, Valdivieso F. Herpes simplex virus type 1 induces nuclear accumulation of hyperphosphorylated tau in neuronal cells. J of Neuroscience Research. 2012;90(5):1020–1029. doi:10.1002/jnr.23003

35. Eimer WA, Vijaya Kumar DK, Navalpur Shanmugam NK, et al. Alzheimer’s Disease-Associated β-Amyloid Is Rapidly Seeded by Herpesviridae to Protect against Brain Infection. Neuron. 2018;99(1):56–63.e3. doi:10.1016/j.neuron.2018.06.030

36. Li H, Liu CC, Zheng H, Huang TY. Amyloid, tau, pathogen infection and antimicrobial protection in Alzheimer’s disease –conformist, nonconformist, and realistic prospects for AD pathogenesis. Transl Neurodegener. 2018;7(1):34. doi:10.1186/s40035-018-0139-3

37. Benn CS, Fisker AB, Rieckmann A, Sørup S, Aaby P. Vaccinology: time to change the paradigm? The Lancet Infectious Diseases. 2020;20(10):e274–e283. doi:10.1016/S1473-3099(19)30742-X

38. Aaby P, Benn CS, Flanagan KL, et al. The non-specific and sex-differential effects of vaccines. Nat Rev Immunol. 2020;20(8):464–470. doi:10.1038/s41577-020-0338-x

