## Supplementary Material for "Association between recombinant herpes zoster vaccination and dementia risk in older adults newly admitted to post-acute and long-term care"

**eFigure 1.** Study design figure.

**eTable 1.** Description of the target trial for estimating the effect of RZV on dementia risk in nursing home residents.

**eTable 2.** Operational definitions of exclusion criteria, exposures, outcomes, and covariates.

**eTable 3.** Cohort flow diagram and exclusions.

**eTable 4.** Distribution of reasons for censoring during the 1-year grace period overall and by treatment assignment.

**eTable 5.** All measured demographic and clinical characteristics of nursing home residents eligible for the target trial at admission and at one year following admission.

**eTable 6.** Model summary of baseline and time-varying covariates associated with receiving the RZV vaccine over time.

**eTable 7.** Distribution of the inverse probability of censor weights for unadjusted, baseline adjusted, and baseline and time-varying adjusted models of vaccination.

**eTable 8.** Cumulative incidence of dementia over time in residents who received at least one dose of RZV within one year of admission compared to no RZV.

**eTable 9.** Absolute and relative risk of dementia between residents who received at least one dose of RZV within one year of admission compared to no RZV at four years of follow-up for all dementia definitions.

**eTable 10.** Absolute and relative risk of negative control outcomes between residents who received at least one dose of RZV within one year of admission compared to no RZV at four years of follow-up.

**eTable 11.** Sensitivity Analyses: absolute and relative risk of outcomes between residents who received at least one dose of RZV within one year of admission compared to no RZV at four years of follow-up.

**eMethods 1.** Overview and results of Medicare data and data linkage for PointClickCare electronic health records and Medicare files.

- **eTable 12.** Description and comparison of nursing home residents who were linked vs. unlinked to Medicare claims.

**eMethods 2.** Clone Censor weight methods.

**eFigure 1.** Study design figure.

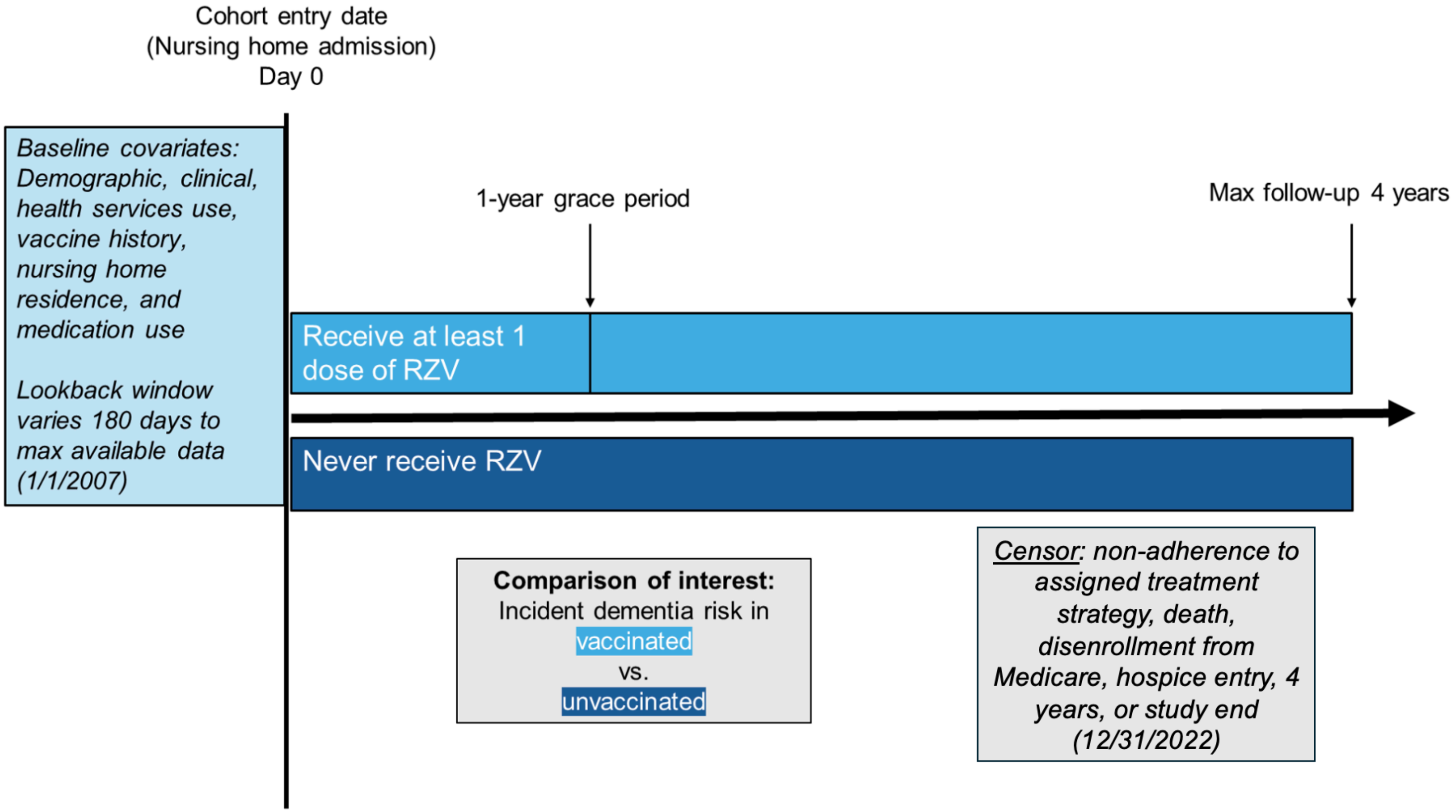

**eTable 1.** Description of the target trial for estimating the effect of RZV on dementia risk in nursing home residents.

| | **Target Trial** | **Emulation in Observational Data** |
|---|---|---|
| **Aim** | Estimate and compare risk of dementia at 1 through 4 years under two recombinant HZ vaccine (RZV) treatment strategies: RZV vaccination within 12 months of nursing home admission or no RZV vaccination through follow-up | |
| **Eligibility** | **Inclusion**<br>● New nursing home residents admitted between January 1, 2017 and December, 31, 2022<br>● Aged 66 years or older as of admission<br>● Has no prior RZV<br><br>**Exclusion**<br>● A baseline history of dementia<br>● A recent history (prior 6 months) of HZ infection<br>● Receipt of hospice/palliative care in the last 6 months and/or upon admission to the facility. | **Inclusion:** Same as the target trial and in addition:<br>● Successfully linked EHR data to Medicare claims files<br>● Enrolled in Medicare Parts A, B, and D for at least 12 months prior to the admission assessment<br><br>**Exclusion**<br>● Same as the target trial<br>● Zero days of follow-up (e.g., died or diagnosed with dementia on the study index date) |
| **Treatment strategies** | (1) Vaccination with at least one dose of RZV within 12 months of nursing home admission,<br><br>(2) No receipt of RZV during follow-up | Same as target trial |
| **Assignment** | Random assignment | Eligible residents are 'cloned' (i.e., baseline data and relevant follow-up events are duplicated in the database) and assigned to both strategies (i.e., one clone assigned to receive the RZV vaccine treatment and the other clone assigned to not receive the vaccine treatment) |
| **Covariates and confounding** | Assumed to be balanced through randomization; balance at baseline is assessed with standardized differences; Marginal effects are estimated for intent-to-treat effect | Demographic, clinical, health services use, vaccine history, nursing home residence, and medication use are measured at baseline and every 30-days over follow-up to capture time-varying changes.<br><br>Baseline and time-varying covariates are included to account for differential censoring due to treatment strategy non-adherence (i.e., clones are administratively censored when their observed data deviates from their assigned treatment strategy). |
| **Outcomes** | Incident dementia diagnosis ascertained through neuropsychological clinical evaluations and tests. | Incident dementia diagnosis ascertained through administrative claims and diagnosis codes neuropsychological clinical evaluations and tests. |
| **Follow-up** | Followed from admission and treatment assignment until the occurrence of the outcome or up to 4 years after nursing home admission. | Same as target trial except we "assign" the treatment at the date of the admission assessment (vs. the entry date into the facility), and stop follow-up additionally at the end of available data |

| | | |
|---|---|---|
| | | (12/31/2023) |
| **Causal Contrasts** | Intention to treat and per-protocol effects | Observational analogue of per-protocol effects |
| **Statistical analysis** | For intention-to-treat effects, unadjusted analyses.<br><br>For per-protocol effects, artificially censor observations if and when they are non-adherent to the assigned treatment strategy; account for artificial censoring using inverse probability weighting. | Per-protocol effects are estimated using the same approach as in the target trial. |
| **Planned secondary analyses** | Residents with documented, prior Zostavax immunization will be examined as a separate subgroup in secondary analyses.<br><br>Assess effect measure modification by sex. | Same as target trial |

**eTable 2.** Operational definitions of exclusion criteria, exposures, outcomes, and covariates.

| | **Definition / data sources** |
|---|---|
| Exclusion: Dementia Diagnosis | MDS admission assessment<br>I4200 = 1 OR I4800=1<br><br>I8000 where the F00.x, F01.x, F02.x, F03.x, G30.x dementia-related ICD-10 codes are indicated (Any position)<br><br>PCC Medication Administrations or Orders<br>class = “antidementia agent”<br><br>ADRD medications: Aricept (donepezil), Exelon (rivastigmine), Namzaric (donepezil/memantine), Aricpet ODT, Razadyne ER (galantamine), Reminyl (galantamine), Razadyne (galantamine), Cognex (tacrine), Adlarity (donepezil), Namenda, Namenda XR (memantine)<br><br>Identify if an anti-dementia drug overlaps the admission assessment date (inclusive of the admission assessment date)<br><br>PCC Diagnosis file<br>F00.x, F01.x, F02.x, F03.x, G30.x (Any position)<br><br>Medicare claims definition for dementia diagnoses and medications<br>Use the Part A, B and D claims.<br><br>one hospitalization code OR (three physician claims codes at least 30 days apart in a one year period) OR a prescription filled for a dementia medication.<br><br>ICD-10 (F00.x, F01.x, F02.x, F03.x, G30.x)<br><br>Flag individuals who meet the definition for dementia prior to the admission assessment date. |
| Exclusion: HZ diagnosis in prior 6 months | PCC active diagnosis file OR MDS active diagnoses (I8000):<br>B02.0: Zoster encephalitis, B02.1: Zoster meningitis, B02.2: Zoster with other nervous system involvement, B02.3: Zoster ocular disease, B02.7: Disseminated zoster, B02.8: Zoster with other complications, B02.9: Zoster without complications<br><br>Medicare claims (Part A and B claims)<br>Same codes as above |

| | |
|---|---|
| Exclusion: prior RZV | Prior RZV v in the PCC-EHR (immunizations, orders, administrations)<br>Using the immunizations file of the EHR, use free-text searches for “shingrix” and “recombinant HZ” vaccinations. (use only investigator-confirmed names)<br><br>Flag individuals with a RZV dated prior to or on the admission assessment date regardless of whether CONSENT = H or CONSENT = Y.<br><br>Using the orders file of the EHR, flag Shingrix orders dated prior to the admission assessment date.<br><br>Using the administrations file of the EHR flag Shingrix administrations dated prior to the admission assessment.<br><br>Prior RZV vaccine in the Part D claims<br>NDC: 58160-819-12, 58160-823-11, 58160-828-01, 58160-828-03, 58160-829-01 |
| Exclusion: hospice | PCC-EHR MDS<br>M3O0100K1=1 or M3O0100K2 = 1 in admission MDS<br><br>A1800 variable - entry from Hospice = “7”<br><br>Medicare claims<br>Z51.5 in Part A and B claims in the prior six months (inclusive of the index date) |
| Exposure: RZV | Same as exclusion, above |
| Outcome: Incident Dementia, Validated Outcome (Dementia 1) | One Part A claim in the hospital Place of Service code = 21 (Hospital) (any diagnostic position) OR Three Part B claims separated by 30 days; Date of the 3rd claim meeting this criteria = has dementia (any diagnostic position) OR One dementia drug in the eMAR or Part D |
| Outcome: Incident Dementia, Sensitive Outcome (Dementia 2) | At least 1 diagnosis code for dementia in Part A OR<br>At least 1 diagnosis code for dementia in Part B OR<br>Checkbox for dementia in the MDS OR<br>At least 1 diagnosis code for dementia in the active diagnosis file of the EHR OR<br>At least 1 diagnosis code for dementia in the MDS (Section I8000) OR<br>At least 1 diagnosis code in the MDS and checkbox for dementia or active diagnosis file PCC OR<br>At least 1 dementia drug in Part D OR<br>At least 1 dementia drug in the eMAR |
| Outcome: Incident Dementia, Specific Outcome (Dementia 3) | At least 1 dementia drug in the eMAR or<br>At least 1 dementia drug in the Part D claims |
| Outcome: Incident HZ | Same as exclusion, above |
| Sensitivity Analysis Outcome: Hip Fracture (Negative control) | Hospitalization (MedPAR) with ICD-10 codes in any position:<br>S72.0 (Fracture of head and neck of femur)<br><br>S72.1 (Pertrochanteric fracture)<br><br>S72.2 (Subtrochanteric fracture of femur) |

| | |
|---|---|
| Sensitivity Analysis Outcome: Wellness Visit (Negative control) | Part B claims data (claim and line files) with the following ICD-10 or CPT/HCPCS codes:<br>ICD-10-CM codes: Z00.00 (Encounter for general adult medical examination without abnormal findings)<br>CPT/HCPCs:<br>G0402 Initial Preventive Physical Examination<br>G0438 for the initial visit<br>G0439 for subsequent visits |
| **Covariates** | |
| Age | MBSF, at index |
| Sex | MBSF, at index |
| Race / Ethnicity | MBSF, at index |
| Region | MBSF or PCC, at index |
| Prior live attenuated HZ vaccine | Prior RZV vaccine in the PCC-EHR (immunizations, orders, administrations), using the same methods as for prior RZV, above OR Part D claim with NDC: 0006-4107-00, 0006-4107-4, in all available data before index (to January 1, 2007) |
| Prior HZ diagnosis<br>(>6 months before index) | Same methods as exclusion, above |
| Cognitive Function Scale | MDS admission assessment (baseline) and follow-up assessments (time-varying) |
| Clinical Diagnoses (e.g., prior myocardial infarction) | MDS admission assessment (baseline) and follow-up assessments (time-varying) and CCW (baseline and time-varying, using the “ever” diagnosed flag that uses all available history) |
| Medications | Part D claims, PCC eMAR records, or MDS assessment (as applicable) in the 30 days before admission, updated over time (ever-use) |
| Vaccinations (COVID-19, flu, pneumonia) | Part D claims, PCC eMAR records, PCC immunizations file in the prior 365 days (baseline) and updated over time (time-varying) |
| Live HZ vaccination | Part D claims, PCC eMAR records, PCC immunizations file in all available data prior to index date (baseline) and updated over time (time-varying) |
| Healthcare utilization (e.g., number of prior hospitalizations) | MedPAR files; Part B Carrier claims in the prior 365 days and updated over time (time-varying) |
| Nursing home discharge | Use MDS tracking records to flag individuals who discharge from the nursing home and do not return within 10 days (time-varying) |

Notes: CPT - Current Procedural Terminology; ICD-10-CM – International Classifications for Diseases, 10$^{th}$ edition, Clinical Modification; eMAR – electronic medication administration record; MBSF - Medicare Beneficiary Summary

| |
|---|
| File; MDS – Minimum Data Set; MedPAR - Medicare Provider Analysis and Review; HZ - herpes zoster; PCC – PointClickCare (electronic health record data); RZV - recombinant HZ vaccine; CCW - Chronic Conditions Warehouse. |

**eTable 3.** Cohort flow diagram and exclusions.

| **Sequential exclusions applied as of the study index date** | N Excluded | Remaining N | % Of Starting N Excluded |
|---|---|---|---|
| Starting N of nursing home admissions linked to Medicare claims | - | 2,667,337 | - |
| < 66 years of age | 377,369 | 2,289,968 | 14.15 |
| < 12 months of continuous enrollment in FFS | 1,062,629 | 1,227,339 | 46.40 |
| < 12 months of continuous enrollment in Part D | 376,216 | 851,123 | 30.65 |
| Has a diagnosis of dementia in the EHR or Medicare claims | 268,847 | 582,276 | 31.59 |
| Receiving hospice care | 31,728 | 550,548 | 5.45 |
| HZ diagnosis as of the past six months in the EHR or Medicare claims | 5717 | 544,831 | 1.04 |
| Has evidence of a prior RZV in the EHR or Medicare claims | 32,436 | 512,395 | 5.95 |
| who had admission date <= 31Dec2022 but admission assessment is greater than 31Dec2022 | 1,417 | 510,978 | 0.28 |
| Zero follow-up time (e.g., dead as of the study index date) | 1,052 | 509,926 | 0.21 |

**eTable 4.** Distribution of reasons for censoring during the 1-year grace period by treatment assignment.

| Characteristic | Assigned to receive 1+ RZV at 1 Year | Not assigned to receive 1+ RZV at 1 year |
|---|---|---|
| Uncensored | 8843 (1.73) | 259518 (50.89) |
| Diagnosed with dementia | 36866 (7.23) | 36685 (7.19) |
| Deviated from assigned treatment strategy | 259369 (50.86) | 10410 (2.04) |
| **Natural censoring** | | |
| Death | 60843 (11.93) | 60669 (11.90) |
| Hospice entry | 53774 (10.55) | 53604 (10.51) |
| Medicare disenrollment | 24486 (4.80) | 24177 (4.74) |
| End of data [December 31, 2022] | 65745 (12.89) | 64863 (12.72) |

**eTable 5.** All measured demographic and clinical characteristics of nursing home residents eligible for the target trial at admission and at one year following admission.

| Characteristic | Overall Cohort | Received 1+ RZV at 1 Year | Did not receive 1+ RZV at 1 year |
|---|---|---|---|
| Total N | **509,926** | **8,843** | **259,518** |
| **Demographic characteristics at baseline** | | | |
| Age, mean (SD) | 79.32 (8.16) | 77.69 (7.13) | 78.74 (7.98) |
| ***Age, n (%)*** | | | |
| 66-69 | 67178 (13.17) | 1251 (14.15) | 36851 (14.20) |
| 70-74 | 98686 (19.35) | 2008 (22.71) | 53427 (20.59) |
| 75-79 | 103771 (20.35) | 2151 (24.32) | 54280 (20.92) |
| 80-84 | 96433 (18.91) | 1768 (19.99) | 48507 (18.69) |
| 85-89 | 78278 (15.35) | 1139 (12.88) | 37854 (14.59) |
| 90+ | 65580 (12.86) | 526 (5.95) | 28599 (11.02) |
| ***Sex, n (%)*** | | | |
| Male | 185471 (36.37) | 2805 (31.71) | 85878 (33.09) |
| Female | 324455 (63.63) | 6038 (68.28) | 173640 (66.91) |
| ***Race and ethnicity, n (%)*** | | | |
| Non-Hispanic White | 430856 (84.49) | 7738 (87.50) | 221520 (85.36) |
| Black | 34711 (6.91) | 302 (3.42) | 16004 (6.17) |
| Other race or ethnicity | 3138 (0.61) | 53 (0.60) | 1611 (0.62) |
| Asian or Pacific Islander | 10959 (2.15) | 274 (3.10) | 5494 (2.12) |
| Hispanic | 23234 (4.56) | 307 (3.47) | 11293 (4.35) |
| American Indian or Alaska Native | 2807 (0.55) | 43 (0.49) | 1438 (0.55) |
| Unknown or missing race or ethnicity | 4221 (0.83) | 126 (1.42) | 2158 (0.83) |
| ***US Geographic Region, n (%)*** | | | |
| Northeast | 108399 (21.26) | 1773 (20.05) | 57079 (21.99) |
| Midwest | 127690 (25.04) | 2352 (26.60) | 65662 (25.30) |
| South | 145016 (28.44) | 2128 (24.06) | 71555 (27.57) |
| West | 111493 (21.86) | 2320 (26.24) | 56663 (21.83) |
| Unknown or missing region | 17328 (3.40) | 270 (3.05) | 8559 (3.30) |
| **Clinical and health services use characteristics at baseline** | | | |

| | | | |
|---|---|---|---|
| Arthritis, n (%) | 421013 (82.56) | 7672 (86.76) | 215811 (83.16) |
| Anemia, n (%) | 429504 (84.23) | 7186 (81.26) | 215076 (82.88) |
| Atrial fibrillation, n (%) | 213349 (41.84) | 3033 (34.30) | 96751 (37.28) |
| Cancer, n (%) | 145402 (28.51) | 2268 (25.65) | 65891 (25.39) |
| Coronary artery disease, n (%) | 117294 (23.00) | 1696 (19.18) | 53870 (20.76) |
| Deep vein thrombosis, n (%) | 19423 (3.81) | 287 (3.25) | 9260 (3.57) |
| Diabetes, n (%) | 280514 (55.01) | 4194 (47.43) | 136562 (52.62) |
| Heart disease (non-ischemic and ischemic), n (%) | 286197 (56.13) | 3849 (43.53) | 132276 (50.97) |
| Herpes zoster, n (%) | 7603 (1.49) | 151 (1.71) | 3248 (1.25) |
| High cholesterol, n (%) | 461403 (90.48) | 8044 (90.96) | 233270 (89.89) |
| Hypertension, n (%) | 401894 (78.81) | 8235 (93.12) | 246302 (94.91) |
| Mental health conditions, n (%) | 300118 (58.86) | 5163 (58.39) | 150124 (57.85) |
| Osteoporosis and fractures, n (%) | 238309 (46.73) | 4497 (50.85) | 125126 (48.21) |
| Parkinson's disease, n (%) | 20485 (4.02) | 270 (3.05) | 9217 (3.55) |
| Peripheral vascular disease, n (%) | 234162 (45.92) | 3307 (37.40) | 109621 (42.24) |
| Renal failure, n (%) | 341588 (66.99) | 5083 (57.48) | 163069 (62.84) |
| Stroke and CVAE, n (%) | 181227 (35.54) | 2455(27.76) | 84189 (32.44) |
| ***Cognitive function scale score, n (%)*** | | | |
| 1 | 359354 (70.47) | 7273 (83.38) | 199865 (77.01) |
| 2 | 104724 (20.54) | 1115 (12.61) | 43742 (16.86) |
| 3 | 29503 (5.79) | 161 (1.82) | 9579 (3.69) |
| 4 | 3676 (0.72) | 20 (0.23) | 992 (0.38) |
| Unknown or missing | 12669 (2.48) | 174 (1.97) | 5340 (2.06) |
| Gagne comorbidity index, mean (SD) | 6.12 (3.60) | 4.61 (3.14) | 5.24 (3.31) |
| **Year of admission to the nursing home, n (%)** | | | |
| 2017 | 85416 (16.75) | 724 (8.19) | 53852 (20.75) |
| 2018 | 91809 (18.00) | 2140 (24.20) | 57500 (22.16) |
| 2019 | 94265 (18.49) | 2927 (33.10) | 57876 (22.30) |
| 2020 | 69664 (13.66) | 1266 (14.32) | 40880 (15.75) |
| 2021 | 82863 (16.25) | 1770 (20.02) | 48918 (18.85) |
| 2022 | 85909 (16.85) | 16 (0.18) | 492 (0.19) |

| | | | |
|---|---|---|---|
| **In NH for 12-month follow-up** | | 1066 (12.05) | 30898 (11.91) |
| **Vaccinations at baseline** | | | |
| COVID-19 vaccine, n (%) | 96715 (18.97) | 1145 (12.95) | 26778 (10.32) |
| Influenza vaccine, n (%) | 216824 (42.52) | 4457 (50.40) | 111155 (42.83) |
| Live HZ vaccine, n (%) | 37822 (7.42) | 1189 (13.45) | 20485 (7.89) |
| Pneumonia vaccine, n (%) | 42781 (8.39) | 855 (9.67) | 23908 (9.21) |
| **Medication use at baseline** | | | |
| Analgesics, n (%) | 262326 (51.44) | 5475 (61.91) | 140894 (54.29) |
| Anticoagulants, n (%) | 221340 (43.41) | 3946 (44.62) | 112723 (43.44) |
| Anticonvulsants, n (%) | 33468 (6.56) | 568 (6.42) | 16025 (6.17) |
| Antidepressant, n (%) | 181860 (35.66) | 3356 (37.95) | 90637 (34.93) |
| Antipsychotic, n (%) | 28906 (5.67) | 465 (5.26) | 13505 (5.20) |
| Antiviral, n (%) | 6752 (1.32) | 163 (1.84) | 3088 (1.19) |
| Benzodiazepine, n (%) | 91042 (17.85) | 1736 (19.63) | 47226 (18.20) |
| Gabapentinoids, n (%) | 103487 (20.29) | 2058 (23.27) | 52429 (20.20) |
| Insulin, n (%) | 104427(20.48) | 1378 (15.58) | 49939 (19.24) |
| **Health services characteristics at baseline** | | | |
| Count of ED visits in the prior year, n (%) | | | |
| 0 | 98947 (19.40) | 1632 (18.46) | 31436 (12.11) |
| 1-4 | 339242 (66.53) | 6336 (71.65) | 195246 (75.23) |
| 5-9 | 61230 (12.01) | 772 (8.73) | 28731 (11.07) |
| 10+ | 10507 (2.06) | 103 (1.16) | 4105 (1.58) |
| Count of hospital visits in the prior year, n (%) | | | |
| 0 | 30421 (5.97) | 470 (5.31) | 14441 (5.56) |
| 1-4 | 454525 (89.14) | 8151 (92.17) | 236242 (91.03) |
| 5-9 | 23688 (4.65) | 211 (2.39) | 8488 (3.27) |
| 10+ | 1292 (0.25) | 11 (0.12) | 347 (0.13) |

**eTable 6.** Model summary of baseline and time-varying covariates associated with receiving the RZV vaccine over time.

| Characteristic | Coefficient | Odds ratio | Z-score |
|---|---|---|---|
| **Demographic characteristics at baseline** | | | |
| Age | -705.737 | <0.000 | -24.620 |
| Age squared | -372.479 | <0.000 | -14.409 |
| Female sex | 0.000 | 1.000 | -0.016 |
| ***Race and ethnicity (referent = Unknown/Missing)*** | | | |
| Non-Hispanic White | -0.410 | 0.664 | -6.914 |
| Black | -0.755 | 0.470 | -10.915 |
| Other race or ethnicity | -0.460 | 0.632 | -4.368 |
| Asian or Pacific Islander | -0.816 | 0.830 | -2.573 |
| Hispanic | -0.572 | 0.564 | -8.231 |
| American Indian or Alaska Native | -0.495 | 0.610 | -4.359 |
| Unknown or missing race or ethnicity | | | |
| ***US Geographic Region (referent = Midwest)*** | | | |
| Northeast | -0.094 | 0.910 | -4.628 |
| South | -0.145 | 0.865 | -7.510 |
| West | 0.070 | 1.073 | 3.605 |
| Unknown or missing region | -0.038 | 0.963 | -0.921 |
| **Clinical and health services use characteristics at baseline** | | | |
| Anemia | 0.018 | 1.019 | 0.987 |
| Arthritis | 0.222 | 1.249 | 10.439 |
| Atrial fibrillation | 0.017 | 1.017 | 1.006 |
| Cancer | 0.052 | 1.053 | 2.844 |
| Coronary artery disease | -0.024 | 0.976 | 1.311 |
| Deep vein thrombosis | -0.158 | 0.854 | -2.18 |
| Diabetes | -0.095 | 0.909 | -5.199 |
| Heart disease (non-ischemic and ischemic) | -0.094 | 0.910 | -5.790 |
| Hypertension | -0.129 | 0.879 | -4.357 |
| High cholesterol | 0.197 | 1.218 | 7.783 |

| | | | |
|---|---|---|---|
| Mental health conditions | -0.047 | 0.954 | -2.674 |
| Osteoporosis and fractures | 0.111 | 1.117 | 7.232 |
| Parkinson's disease | -0.179 | 0.836 | -4.558 |
| Peripheral vascular disease | -0.031 | 0.970 | -2.001 |
| Renal failure | -0.020 | 0.980 | -1.181 |
| Stroke and CVAE | -0.052 | 0.949 | -3.120 |
| Prior non-hip fracture | -0.028 | 0.973 | -0.570 |
| ***Cognitive function scale score (referent = 1)*** | | | |
| 2 | -0.239 | 0.787 | -11.375 |
| 3 | -0.486 | 0.615 | -10.183 |
| 4 (severe cognitive impairment) | -0.203 | 0.816 | -1.617 |
| Unknown or missing CFS | 0.023 | 1.023 | 0.474 |
| ***Activities of daily living scale score (referent = ≥15)*** | | | |
| 0-4 (no to few limitations) | 0.398 | 1.488 | 6.077 |
| 5-9 | 0.29 | 1.336 | 4.415 |
| 10-14 | 0.122 | 1.130 | 1.847 |
| Unknown or missing ADL | 0.323 | 1.381 | 4.732 |
| ***Gagne comorbidity index (referent = 0)*** | | | |
| 1-4 | 0.272 | 1.313 | 1.204 |
| 5-9 | 0.237 | 1.268 | 1.045 |
| 10-14 | 0.120 | 1.128 | 0.526 |
| 15-19 | -0.026 | 0.975 | -0.104 |
| 20+ (substantial comorbidity) | -1.147 | 0.317 | -1.118 |
| ***Emergency department visits as of the prior 365 days (referent = 1-4)*** | | | |
| 0 | 0.219 | 1.244 | 11.341 |
| 5-9 | -0.120 | 0.887 | -4.591 |
| 10+ | -0.153 | 0.858 | -2.207 |
| Part B claim in the prior 365 days | 0.051 | 0.950 | -2.753 |
| **Year of admission to the nursing home (referent = 2017)** | | | |
| 2018 | 0.484 | 1.623 | 21.021 |
| 2019 | 0.629 | 1.876 | 25.797 |

| | | | |
|---|---|---|---|
| 2020 | 0.117 | 1.124 | 3.778 |
| 2021 | 0.344 | 1.410 | 9.296 |
| 2022 | 0.056 | 1.058 | 1.194 |
| **Month of admission to the nursing home (referent = 1)** | | | |
| 2 | -0.020 | 0.980 | -0.626 |
| 3 | -0.006 | 0.994 | -0.183 |
| 4 | -0.069 | 0.933 | -2.096 |
| 5 | -0.012 | 0.988 | -0.38 |
| 6 | -0.003 | 0.997 | -0.098 |
| 7 | -0.087 | 0.917 | -2.588 |
| 8 | -0.036 | 0.965 | -1.083 |
| 9 | -0.007 | 0.993 | -0.221 |
| 10 | -0.07 | 0.932 | -2.077 |
| 11 | -0.064 | 0.938 | -1.911 |
| 12 | 0.005 | 1.005 | 0.150 |
| **Medication and vaccinations at baseline** | | | |
| Flu vaccine | 0.352 | 1.423 | 22.392 |
| COVID-19 vaccine | 0.165 | 1.179 | 4.813 |
| Pneumonia vaccine | 0.029 | 1.029 | 1.263 |
| Zostavax vaccine | 0.629 | 1.875 | 31.828 |
| Analgesic | 0.070 | 1.073 | 4.618 |
| Anticoagulants | 0.07 | 1.072 | 4.672 |
| Anticonvulsant | 0.060 | 1.062 | 2.13 |
| Antidepressant | 0.105 | 1.111 | 5.997 |
| Antipsychotic | 0.028 | 1.028 | 0.869 |
| Antiviral | 0.222 | 1.249 | 4.100 |
| Benzodiazepine | 0.040 | 1.041 | 2.220 |
| Gabapentinoids | 0.104 | 1.109 | 6.140 |
| Insulin | -0.152 | 0.859 | -2.766 |
| **Time-varying clinical and health services use characteristics** | | | |
| Diabetes | 0.054 | 1.056 | 2.608 |

| | | | |
|---|---|---|---|
| Herpes zoster diagnosis | 0.784 | 2.190 | 16.827 |
| Stroke and CVAE | -0.050 | 0.952 | -2.317 |
| Deep vein thrombosis | 0.103 | 1.109 | 1.631 |
| Nursing home discharge | 0.199 | 1.220 | 9.350 |
| ***Emergency department visits (referent = 1-4)*** | | | |
| 0 | 0.158 | 1.171 | 4.81 |
| 5-9 | -1.098 | 0.334 | -1.551 |
| 10+ | -5.794 | 0.003 | -0.178 |
| ***Hospitalizations (referent = 1-4)*** | | | |
| 0 | 0.654 | 1.924 | 16.862 |
| 5-9 | -5.197 | 0.006 | -0.183 |
| **Time-varying medication and vaccinations** | | | |
| COVID-19 vaccine | 0.382 | 1.465 | 16.219 |
| Flu vaccine | 0.273 | 1.314 | 15.326 |
| Pneumonia vaccine | 0.395 | 1.485 | 12.262 |
| Zostavax vaccine | 0.567 | 1.762 | 4.051 |
| Anticoagulants | -0.273 | 0.761 | -4.830 |
| Antidepressant | -0.248 | 0.780 | -4.255 |
| Antipsychotic | -0.003 | 0.997 | -0.026 |
| Antiviral | 0.007 | 1.007 | 0.032 |
| Insulin | 0.085 | 1.008 | 1.514 |

**eTable 7.** Distribution of the inverse probability of censor weights for unadjusted, baseline adjusted, and baseline and time-varying adjusted models of vaccination.

| **Treatment Arm and Model Specification** | **Minimum** | **Maximum** | **Mean** | **Median** | **90th percentile** | **95th percentile** | **99th percentile** |
|---|---|---|---|---|---|---|---|
| *Received 1+ RZV at 1 Year* | | | | | | | |
| Unadjusted | 1.00 | 37.7 | 1.15 | 1.00 | 1.00 | 1.00 | 1.00 |
| Baseline adjusted | 1.00 | 162 | 1.14 | 1.00 | 1.00 | 1.00 | 1.00 |
| Full adjusted | 1.00 | 161.00 | 1.13 | 1.00 | 1.00 | 1.00 | 1.00 |
| *Did not receive 1+ RZV at 1 Year* | | | | | | | |
| Unadjusted | 1.00 | 1.10 | 1.04 | 1.03 | 1.08 | 1.09 | 1.10 |
| Baseline adjusted | 1.00 | 2.03 | 1.04 | 1.03 | 1.09 | 1.13 | 1.22 |
| Full adjusted | 1.00 | 3.56 | 1.04 | 1.03 | 1.10 | 1.13 | 1.22 |

**eTable 8.** Cumulative incidence of dementia over time in residents who received at least one dose of RZV within one year of admission compared to no RZV.

| | Cumulative number of events: 1+ RZV | Cumulative number of events: No RZV | Cumulative incidence: 1+ RZV | Cumulative incidence: No RZV | Absolute Risk Difference (95%CI) | Risk Ratio (95%CI) |
|---|---|---|---|---|---|---|
| **Primary, Validated Dementia Outcome** | | | | | | |
| Year 1 | 41,217 | 41,026 | 10.00% | 10.46% | -0.46% (-0.60%, -0.33%) | 0.96 (0.94, 0.97) |
| Year 2 | 41,460 | 54,293 | 13.61% | 15.76% | -2.15% (-2.66%, -1.65%) | 0.86 (0.83, 0.90) |
| Year 3 | 41,650 | 61,743 | 15.99% | 19.86% | -3.87% (-4.79%, -2.88%) | 0.81 (0.76, 0.85) |
| Year 4 | 41,736 | 65,913 | 18.75% | 24.56% | -5.81% (-7.47%, -3.85%) | 0.76 (0.69, 0.84) |
| **Sensitive Dementia Outcome Definition** | | | | | | |
| Year 1 | 44,150 | 43,948 | 10.64% | 11.21% | -0.57% (-0.72%, -0.42%) | 0.95 (0.94, 0.96) |
| Year 2 | 44,400 | 57,268 | 14.03% | 16.44% | -2.41% (-2.90%, -1.92%) | 0.85 (0.82, 0.88) |
| Year 3 | 44,588 | 64,716 | 16.32% | 20.41% | -4.10% (-4.92%, -3.23%) | 0.80 (0.76, 0.84) |
| Year 4 | 44,682 | 68,829 | 19.46% | 25.16% | -5.71% (-7.47%, -3.96%) | 0.77 (0.70, 0.84) |
| **Specific Dementia Outcome Definition (Medication Use)** | | | | | | |
| Year 1 | 9,802 | 9,726 | 2.55% | 2.48% | 0.07% (-0.02%, 0.16%) | 1.03 (0.99, 1.07) |
| Year 2 | 9,895 | 12,958 | 4.02% | 3.94% | 0.09% (-0.28%, 0.51%) | 1.02 (0.93, 1.13) |
| Year 3 | 9,955 | 14,766 | 5.06% | 5.00% | -0.06% (-0.72%, 0.74%) | 0.99 (0.86, 1.14) |
| Year 4 | 9,983 | 15,693 | 5.73% | 6.18% | -0.45% (-1.47%, 0.70%) | 0.93 (0.76, 1.11) |

**eTable 9.** Absolute and relative risk of dementia between residents who received at least one dose of RZV within one year of admission compared to no RZV at four years of follow-up for all dementia definitions.

| Model | Cumulative incidence of dementia at 4 years: 1+ RZV | Cumulative incidence of dementia at 4 years: No RZV | Absolute Risk Difference (95%CI) | Risk Ratio (95%CI) |
|---|---|---|---|---|
| **Primary, Validated Dementia Outcome** | | | | |
| Unadjusted | 18.32% | 24.78% | -6.46% (-8.05%, -4.73%) | 0.74 (0.66, 0.81) |
| Baseline Adjusted | 19.04% | 24.57% | -5.53% (-7.38%, -3.39%) | 0.77 (0.70, 0.86) |
| Full adjusted | 18.75% | 24.56% | -5.81% (-7.47%, -3.85%) | 0.76 (0.69, 0.84) |
| **Sensitive Dementia Outcome Definition** | | | | |
| Unadjusted | 19.26% | 25.39% | -6.13% (-7.82%, -4.26%) | 0.76 (0.69, 0.83) |
| Baseline Adjusted | 19.77% | 25.17% | -5.39% (-7.41%, -3.39%) | 0.79 (0.71, 0.86) |
| Full adjusted | 19.46% | 25.16% | -5.71% (-7.47%, -3.96%) | 0.77 (0.70, 0.84) |
| **Specific Dementia Outcome Definition (Medication Use)** | | | | |
| Unadjusted | 5.84% | 6.20% | -0.36% (-1.42%, 0.89%) | 0.94 (0.77, 1.14) |
| Baseline Adjusted | 5.80% | 6.18% | -0.37% (-1.43%, 0.89%) | 0.94 (0.77, 1.14) |
| Full adjusted | 5.73% | 6.18% | -0.45% (-1.47%, 0.70%) | 0.93 (0.76, 1.11) |

**eTable 10.** Absolute and relative risk of negative control outcomes between residents who received at least one dose of RZV within one year of admission compared to no RZV at four years of follow-up.

| | Cumulative incidence at 4 years: 1+ RZV | Cumulative incidence at 4 years: No RZV | Absolute Risk Difference (95%CI) | Risk Ratio (95%CI) |
|---|---|---|---|---|
| **Hip fracture (acute)** | | | | |
| Unadjusted | 5.50% | 6.80% | -1.30% (-2.30%, -0.07%) | 0.81 (0.66, 0.99) |
| Baseline Adjusted | 5.74% | 6.77% | -1.02% (-2.12%, 0.35%) | 0.85 (0.69, 1.05) |
| Full adjusted | 5.80% | 6.77% | 0.97% (-2.10%, 0.47%) | 0.86 (0.69, 1.07) |
| **Wellness visit** | | | | |
| Unadjusted | 34.62% | 30.46% | 4.16% (1.69%, 6.43%) | 1.14 (1.06, 1.21) |
| Baseline Adjusted | 33.11% | 30.53% | 2.58% (0.22%, 4.86%) | 1.08 (1.01, 1.16) |
| Full adjusted | 33.06% | 30.56% | 2.50% (0.13%, 4.62%) | 1.08 (1.00, 1.15) |

**eTable 11.** Sensitivity analyses: absolute and relative risk of outcomes between residents who received at least one dose of RZV within one year of admission compared to no RZV at four years of follow-up.

| Model | Cumulative incidence at 4 years: 1+ RZV | Cumulative incidence at 4 years: No RZV | Absolute Risk Difference (95%CI) | Risk Ratio (95%CI) |
|---|---|---|---|---|
| **Competing Risk of Death** | | | | |
| Dementia | 18.71% | 24.51% | -5.79% (-5.93%, -5.65%) | 0.76 (0.76, 0.77) |
| Death | 24.80% | 32.84% | -8.04% (-8.23%, -7.87%) | 0.76 (0.75, 0.76) |
| **Censoring on NH discharge** | | | | |
| Dementia | 18.45% | 24.96% | -6.51% (-10.80%, -2.31%) | 0.74 (0.57, 0.91) |
| **IPCW and modeling modifications** | | | | |
| Weights truncated at the 99th percentile | 18.63% | 24.58% | -5.95% (-6.59%, -5.28%) | 0.76 (0.73, 0.78) |
| Stabilized weights | 18.68% | 24.56% | -5.88% (-6.56%, -5.16%) | 0.76 (0.73, 0.79) |
| Baseline covariates included in outcome model | 12.15% | 19.60% | -7.45% (-8.84%, -5.86%) | 0.62 (0.55, 0.70) |

**eMethods 1.** Overview and results of Medicare data and data linkage for PointClickCare electronic health records and Medicare files.

*Medicare Datasets*

Medicare data included the Medicare Beneficiary Summary File (MBSF) containing demographics, enrollment, and Chronic Conditions Warehouse diagnoses (e.g., hypertension, diabetes, heart failure); Medicare Provider Analysis and Review (MedPAR) data, containing claims and relevant diagnoses from hospitalizations and skilled nursing facilities; Medicare Part B carrier claims for outpatient and emergency department visits; and Medicare Part D claims for prescription medication dispensings and select vaccinations.

*Linkage Methods*

Residents residing in a nursing home (NH) facility using PointClickCare(R) as their electronic health record were linked deterministically to Medicare data using three identifiers: social security number (SSN), health insurance number (HIN), and Medicare beneficiary number (MBN). We restricted the population to residents with a strong and unambiguous match to a Medicare beneficiary, defined as 1) matching SSN, HIN, or MBN; and 2) matching sex; and 3) matching date of birth; and 4) only one potential match based on criteria 1-3. General Dynamics Information Technology (contractor of the Centers for Medicare & Medicaid Services) created a crosswalk of unique and encrypted identifiers to link eligible Medicare beneficiaries to the NH EHR records.

*Linkage Results*

Of the 3,563,893 PCC residents with a new admission during the study period in the EHR data, 2,667,337 (75%) had a “strong” match to Medicare. On average, those with a strong match vs. no match/weak match were older (mean age 77.0 vs. 69.7 years and more likely to be female (58% vs. 54%; **eTable 12, below**).

**eTable 12.** Description and comparison of nursing home residents who were linked vs. unlinked to Medicare claims.

| Characteristic | Linked to Medicare claims | Not linked to Medicare claims |
|---|---|---|
| Total N | 2,667,337 | 896,556 |
| **Demographic characteristics at baseline** | | |
| Age, mean (SD) | 77.02 (11.09) | 69.73 (14.76) |
| ***Age, n (%)*** | | |
| 0-66 | 377369 (14.15) | 339438 (37.86) |
| 66-69 | 259845 (9.74) | 77251 (8.62) |
| 70-74 | 401088 (15.04) | 110042 (12.27) |
| 75-79 | 446114 (16.73) | 113168 (12.62) |
| 80-84 | 448895 (16.83) | 104859 (11.70) |
| 85-89 | 400759 (15.02) | 86895 (9.69) |
| 90+ | 333267 (12.49) | 64774 (7.22) |
| Missing | - | 129 (0.01) |
| ***Sex, n (%)*** | | |
| Male | 1116782 (41.87) | 409355 (45.66) |
| Female | 1550555 (58.13) | 485284 (54.13) |
| Missing | - | 1917 (0.21) |
| **Clinical and health services use characteristics at baseline** | | |
| Cancer, n (%) | 271378 (10.17) | 77160 (8.61) |
| Deep vein thrombosis, n (%) | 102973 (3.86) | 43982 (4.91) |
| Parkinson's disease, n (%) | 98909 (3.71) | 30977 (3.46) |
| Hypertension, n (%) | 2041953 (76.55) | 647363 (72.21) |
| ***Cognitive function scale score, n (%)*** | | |
| 1 | 1549849 (58.10) | 552933 (61.67) |
| 2 | 602277 (22.58) | 189791 (21.17) |
| 3 | 378824 (14.20) | 105416 (11.76) |
| 4 | 61451 (2.30) | 21378 (2.38) |
| Unknown or missing CFS | 74936 (2.81) | 27038 (3.02) |
| **Year of admission to the nursing home, n (%)** | | |

| | | |
|---|---|---|
| 2017 | 389890 (14.62) | 105720 (11.79) |
| 2018 | 429628 (16.11) | 133074 (14.84) |
| 2019 | 462318 (17.33) | 158668 (17.70) |
| 2020 | 375099 (14.06) | 138495 (15.45) |
| 2021 | 467593 (17.53) | 176050 (19.64) |
| 2022 | 534090 (20.02) | 182081 (20.31) |
| 2023 | 8719 (0.33) | 2468 (0.28) |

Notes: Exclusions were applied after linkage to Medicare, so excluded populations (e.g., < 66 years of age) will be present in this table.

**eMethods 2.** Clone Censor Weight Methods.

We utilized the "clone-censor-weight" approach in the target trial emulation.[1,2] To reduce immortal time bias, this method defines a clear time zero (i.e., nursing home admission assessment date), and aligns the assessment of study eligibility, treatment assignment, and start of follow-up as of this date.[2] However, because residents' observed data can be consistent with both treatment strategies (i.e., periods of both exposed and unexposed time within the same individual) each person is "cloned" and assigned to a different treatment strategy. Cloning creates two identical records in the dataset for every eligible NH resident. Clones were treated as separate observations and they were artificially censored when they were no longer adherent to their assigned treatment strategy (i.e., emulating the per-protocol estimand of a trial).

The first 12 months of follow-up was considered a "grace period" to allow for clones who were assigned to vaccination to receive at least one dose of RZV. These clones accrued follow-up time (and thus had outcomes measured) for the entire grace period, but only those who received a dose of RZV were followed after the 12-month grace window.

Naive estimates at each point of follow-up are likely biased because artificial censoring is informative (i.e., there are likely common causes of censoring for RZV receipt and dementia). To address this, we estimated the inverse probability of remaining uncensored (IPCW) for each treatment strategy.[3,4] The weighted analysis estimates cumulative probabilities if the population had no censoring for treatment nonadherence. IPCW are estimated separately under each treatment strategy using pooled logistic regression. Since censoring is related to vaccination status across time, the regression models included both time-fixed and time-varying predictors of RZV vaccination. To explore which variables were associated with RZV vaccination, we assessed both odds ratios and Z-scores from the pooled logistic regression models. Pooled logistic regression models with IPCW compared the cumulative incidence of dementia in 30-day intervals between the vaccine treatment groups on the absolute (risk differences) and relative (risk ratio) scales.

### REFERENCES


1. Gaber CE, Ghazarian AA, Strassle PD, et al. De-Mystifying the Clone-Censor-Weight Method for Causal Research Using Observational Data: A Primer for Cancer Researchers. *Cancer Med*. 2024;13(23):e70461. doi:10.1002/cam4.70461

2. Hernán MA, Sauer BC, Hernández-Díaz S, Platt R, Shrier I. Specifying a target trial prevents immortal time bias and other self-inflicted injuries in observational analyses. *J Clin Epidemiol*. 2016;79:70-75. doi:10.1016/j.jclinepi.2016.04.014

3. Zhao SS, Lyu H, Yoshida K. Versatility of the clone-censor-weight approach: response to “trial emulation in the presence of immortal-time bias.” *International Journal of Epidemiology*. 2021;50(2):694-695. doi:10.1093/ije/dyaa223

4. Duchesneau ED, Jackson BE, Webster-Clark M, et al. The Timing, the Treatment, the Question: Comparison of Epidemiologic Approaches to Minimize Immortal Time Bias in Real-World Data Using a Surgical Oncology Example. *Cancer Epidemiol Biomarkers Prev*. 2022;31(11):2079-2086. doi:10.1158/1055-9965.EPI-22-0495
